# Language History Collection in Multilingual Clinical Practice: A Qualitative Analysis of Public-Sector Clinical Perspectives

**DOI:** 10.1101/2025.09.02.25334739

**Authors:** Kai Ian Leung, Robyn Westmacott, Elizabeth Rochon, Monika Molnar

**Author notes:** Correspondence to Kai Ian Leung at Department of Speech-Language Pathology, University of Toronto, 500 University Avenue, Toronto, ON, M5G 1V7, Canada. **Author Note** Kai Ian Leung Robyn Westmacott Elizabeth Rochon Monika Molnar.

## Abstract

**Background:** Clinicians increasingly work with multilingual paediatric clients across healthcare and community settings. Collecting detailed language background is a crucial first step in planning effective assessment and intervention. Yet, little is known about how this process unfolds in everyday public-sector clinical practice. To improve service quality, equity, and effectiveness for multilingual children, this study investigates how clinicians gather, interpret, and use language history information: as well, it examines the institutional and professional barriers and facilitators that shape this aspect of clinical practice.

**Methods:** A qualitative study was conducted using semi-structured interviews with 21 clinicians working in public-sector and community-based settings across Canada. Data was analysed using framework analysis, guided by the Theoretical Domains Framework.

**Results:** Clinicians universally recognized the value of language history and routinely embedded it within the broader case history. However, variability emerged in what information was gathered, how it was elicited, and how it was used. Practices were shaped by clinician experience, institutional processes, documentation systems, and availability of training and tools. Many relied on flexible, conversational strategies over research-developed tools, often constructing their own frameworks in response to contextual demands. This adaptability reflected the development of adaptive expertise but also risked inconsistencies in data quality, especially in the absence of formal guidance, structured tools, or interpreter support.

**Conclusion:** Language history collection is a complex, multidimensional task influenced by clinician initiative and systemic constraints. Strengthening practice will require hybrid tools that balance structure with flexibility, clearer protocols across disciplines, and institutional investments in interpreter services, training, and culturally informed workflows.

**WHAT THIS PAPER ADDS:** *Section 1: What is already known on this subject:* Collecting detailed language history is essential when assessing multilingual children, as it serves as a foundational step in guiding service delivery. Existing research-developed questionnaires (e.g., LEAP-Q, ALDeQ) provide structured ways of gathering this information, but evidence about how clinicians actually collect and use language history in everyday practice is limited.

*Section 2: What this paper adds to existing knowledge:* This study provides qualitative evidence on how public-sector clinicians in Canada gather, interpret, and apply language history information with multilingual children. It shows that while language history is universally valued, practices vary widely depending on clinician experience, institutional systems, and resource availability. Findings highlight both adaptive strategies and systemic gaps, pointing to the need for hybrid approaches that combine structure with flexibility.

*Section 3: What are the potential or actual clinical implications of this work?:* This study showed that while clinicians’ flexible, conversational strategies promote rapport and cultural responsiveness, they may also create variability and leave gaps in completeness and reliability. Clinicians can integrate hybrid approaches that combine structure with adaptability to support gathering more consistent and clinically useful information. At the system level, standardized documentation protocols, access to trained interpreters, and interprofessional training are critical supports necessary for embedding consistent, high-quality language history collection across services.

## Introduction

Growing recognition of linguistic diversity among client populations is beginning to reshape public-sector service delivery worldwide. Speech-language pathologists (SLPs) and related professionals are increasingly called upon to assess and support bi(multi)lingual families whose cultural and/or linguistic experiences may differ significantly from their own (American Speech-Language-Hearing Association, 2023; Royal College of Speech and Language Therapists, 2006; Speech Pathology Australia, 2023; Speech-Language and Audiology Canada, 2024). In this context, the case history—especially the language history or profile—has emerged as a crucial component of culturally responsive and clinically accurate assessment (McLeod et al., 2017; Shipley & McAfee, 2023).

A comprehensive language history (or language profile) explores the patient’s linguistic environment and experiences, encompassing simultaneous and/or subsequent language acquisition, patterns of language use (e.g., interlocutors’ information, school vs home), proficiency across modalities (e.g., written, read, overheard, spoken, signed), developmental changes over time and more (Kašćelan et al., 2022; Leung & Molnar, 2025). Accurate language profiling is a recommended practice in speech-language pathology and psychology (American Psychological Association, 2020; American Speech-Language-Hearing Association, 2023; Fujii, 2018; International Expert Panel on Multilingual Children’s Speech, 2012; Multilingual-Multicultural Affairs Committee, International Association of Communication Sciences and Disorders, 2011, 2020; Speech-Language and Audiology Canada, 2024). Without this information, clinicians risk misinterpreting typical bilingual language patterns as signs of disorder or failing to identify genuine difficulties masked by code-switching or uneven exposure (Kohnert, 2010; Paradis, 2011). Collecting language background information is a foundational first step of the assessment that should guide the rest of the assessment and then intervention planning. Having accurate information about the client’s language background can minimize over- or under-identification of language-related disorders.

To collect language histories, clinicians employ a range of methods—including interviews, questionnaires, observational data, and review of records. Clinical guidelines for working with developmental populations commonly recommend the use of the case history as a reliable means of gathering language history information, usually through parent questionnaires and/or interviews (American Speech-Language-Hearing Association, n.d.; Canadian Association of Social Workers, 2024; Douglas et al., 2014; Speech-Language and Audiology Canada, 2024). Despite the recognized importance of language history in the evaluation of multilingual clients and its reported high use among SLPs (Dubasik & Valdivia, 2020; Hallin & Partanen, 2024; Hernandez et al., 2025; Hunt et al., 2025), relatively little is known about how clinicians actually gather this information in day-to-day practice. Research has emphasized the need for culturally responsive assessment tools, yet the specific procedures and tools used to collect detailed language histories, especially in paediatric populations, remain underdeveloped. For instance, there are few protocols for surveying the clinical history of infants and preschoolers available, with none focusing on language profiling (Melo et al., 2022). Numerous studies have identified both barriers and facilitators to practices surrounding care of culturally and linguistically diverse clients – including limited interpreter access, insufficient training, resource constraints, as well as proposed supports such as enhanced professional development, culturally-appropriate assessment tools, and broader systemic change (Cao & Keller-Bell, 2025; Fulcher-Rood & Castilla-Earls, 2023; Fumero et al., 2021; Guiberson & Atkins, 2012; Hallin & Partanen, 2024; Hernandez et al., 2025; Hunt et al., 2025; Kritikos, 2003; Lal et al., 2023; McKenna et al., 2025; NoorAli et al., 2025; Parveen & Santhanam, 2021; Santhanam et al., 2019; Suswaram et al., 2023; Teoh et al., 2018; Verdon et al., 2015; Williams & McLeod, 2012). However, how these factors specifically influence the collection, interpretation, and application of language history information in clinical practice remains underexplored.

Clinical settings influence how language history is collected. In Canada, where this study was conducted, public-sector SLPs work across schools, hospitals, and community health centres. National data show that hospitals are the second most common workplace after schools, with smaller proportions in community and public health settings (Speech-Language and Audiology Canada, 2018). Similar patterns exist in the U.S., where 42% of ASHA members work in healthcare (American Speech-Language-Hearing Association, 2025). Compared to school-based peers, healthcare SLPs more often use interviews, conversational samples, and collaborate with parents, methods well-suited to medically complex cases where language profiles may intersect with co-occurring conditions (Fulcher-Rood & Castilla-Earls, 2023; Hallin & Partanen, 2024; Selin et al., 2022). In team-based care, SLPs work alongside professionals who also collect linguistic and cultural data, yet standardized protocols for gathering and interpreting this information remain limited.

### The Current Study

This study explores how public-sector clinicians approach the collection of language history information when working with multilingual clients. Focusing on two research questions, the purpose is to understand both the practices in situ of language history collection, with attention to individual and system level factors:

1. How do public-sector clinicians collect, understand and use language history information in their clinical work with multilingual clients?
2. What barriers, facilitators, and motivations influence clinicians’ decisions to collect, interpret, and apply language history information in practice?

The current research was conducted in Canada, where just like in many other countries, a multilingual clientele is common, especially in urban settings. Nearly one in four Canadians speaks a mother tongue other than English or French (Statistics Canada, 2022b), positioning public-sector clinicians at the forefront of culturally responsive care. Comparable multilingual realities are found in other English-dominant language countries such as the United States, the United Kingdom, and Australia (Williams & McLeod, 2012). To date, no studies have examined how public sector clinicians collect language histories in practice, or how contextual and systemic factors influence their approaches. Addressing this gap is essential to improving assessment equity and supporting evidence-based care for multilingual populations.

## Methods

### Study Design and Theoretical Orientation

We conducted a qualitative study using semi-structured interviews with public-sector clinicians to explore their experiences of language history collection. Grounded in a pragmatic paradigm and informed by a pluralist ontology, our design emphasized methodological flexibility and recognized both individual experiences and institutional structures shaping practice (Morgan, 2014; Onwuegbuzie & Leech, 2005). Interview questions were informed by the Theoretical Domains Framework (TDF; Cane et al., 2012), supporting a theoretically grounded exploration of influencing factors. We used framework analysis, an approach suitable for applied health research aiming to inform practice and policy (Gale et al., 2013; Ritchie et al., 1994).

This study received ethics approval from the University of Toronto’s Health Research Ethics Board (Protocol #46019). Reporting follows the Consolidated Criteria for Reporting Qualitative Research (COREQ; Tong et al., 2007).

### Eligibility Criteria and Recruitment

We employed maximal variation sampling to ensure a range of clinical perspectives across publicly funded Canadian settings. Eligible participants were (1) currently employed in the public sector and (2) regularly working with multilingual paediatric client families. We recruited clinicians whose roles intersect with language, including speech-language pathologists, neuropsychologists, clinical psychologists, applied behavioural analysts, nurses, social workers, and physicians. Recruitment occurred via outreach to professional organizations, clinical networks, social media (e.g., Facebook, LinkedIn, X/Twitter), and snowball sampling.

### Data Collection

Interested clinicians completed a screening questionnaire to confirm eligibility, followed by electronic informed consent and a demographic and language background survey. Data were collected through the University of Toronto’s Research Electronic Data Capture platform (Harris et al., 2009, 2019) and included age, gender, profession, location, work setting, and languages spoken. These details supported both sample diversity and interpretation of findings.

Interviews were conducted virtually at a location chosen by each participant and lasted 45–60 minutes. A pilot interview with a neuropsychologist on the research team (RW) helped refine the guide and process but was excluded from analysis. The interview guide (Supplementary Materials) was co-developed by KIL and MM and drew on six TDF domains (Cane et al., 2012; see Supplementary Materials for domain definitions). The guide included four core questions with probes mapped to the TDF. While the same topics were addressed across participants, question order was adapted to sustain conversational flow and explore participant narratives. Field notes were taken during interviews to support facilitation.

Transcripts were generated using noScribe, an AI transcription tool operating locally for data security (v0.5; Dröge, 2024) and manually reviewed for accuracy. Personally identifying information was pseudonymized before analysis by a trained research assistant and KIL.

### Researcher Team and Reflexivity

The research team engaged in reflexive practices to acknowledge positionality and potential sources of bias. The lead interviewer (KIL) is a multilingual, cisgender woman and PhD candidate in speech-language pathology, with lived and academic experience in multilingual contexts. She had no prior relationship with participants, reducing social desirability bias. The broader team brought interdisciplinary expertise across speech-language pathology, neuropsychology, and bilingualism research, as well as diverse lived experiences related to language, culture, and identity. These perspectives informed study design, coding, and interpretation. Reflexivity was maintained through memo writing, analytic dialogue, and regular team debriefs throughout data collection and analysis.

### Analytical Approach

We used framework analysis (Gale et al., 2013; Ritchie et al., 1994), which supports both inductive and deductive theme development and offers structured data management. Analysis followed five stages: *Familiarization* – KIL reviewed all transcripts and open-ended survey responses, recording initial impressions. Selected transcripts were also reviewed by the team. *Identifying a Thematic Framework* – An initial codebook was developed using both inductive codes from participants’ language and deductive codes from the interview guide and TDF domains. The team iteratively refined the framework to resolve overlaps and improve clarity. *Indexing* – The final codebook was applied in NVivo 14 (Lumivero, 2023). KIL and a trained assistant independently coded four transcripts, discussed discrepancies, and refined the framework. Inter-coder reliability was assessed through thematic agreement and matrix comparisons. *Charting* – A framework matrix in NVivo organized coded data by theme and participant, with each cell summarizing content and illustrative quotes. *Mapping and Interpretation* – Themes and subthemes were identified by KIL and reviewed in collaboration with MM. Relationships between interpersonal and structural factors were explored through memoing and team discussion until analytic consensus was reached.

We applied several strategies to enhance trustworthiness, consistent with qualitative research standards (Lincoln & Guba, 1986). Team-based coding ensured that themes reflected diverse interpretations. An audit trail was maintained through versioned codebooks, analytic memos, and decision logs. Peer debriefing among team members supported confirmability. Researchers documented positionality and assumptions throughout the process. While transcript and theme checking with participants was not conducted due to time constraints, representative quotations are included to support transparency and enable credibility assessment by readers.

## Results

In line with the study’s pragmatic approach, interviews were conducted with 21 clinicians from varied practice settings across five Canadian provinces (Ontario, Alberta, British Columbia, Newfoundland & Labrador, Prince Edward Island). Participants were purposively selected to ensure diversity in professional discipline, experience, geography, and language background. Recruitment concluded upon reaching thematic saturation, with sufficient depth and diversity across thematic categories. All participants consented to take part and none withdrew from the study. The sample included speech-language pathologists (57%), neuropsychologists (14%), social workers (14%), nurses (10%), and clinical psychologists (5%). Clinical experience ranged from 1 to over 20 years, with one-third reporting 11–20 years of practice. Most participants were female, and more than half reported bi(multi)lingual proficiency. While English was the primary language of service delivery, care was also provided in French, Italian, Urdu, or Cantonese. Clinicians worked in hospitals (57%), regional health authorities (29%), and community-based services (14%), predominantly in urban areas (76%).

**Table 2.** Demographic, professional, and linguistic characteristics of participating clinicians.

| Characteristic | n | % |
| --- | --- | --- |
| **Profession** |  |  |
| Speech-language pathologist | 12 | 57% |
| Neuropsychologist | 3 | 14% |
| Clinical psychologist | 1 | 5% |
| Social worker | 3 | 14% |
| Nurse | 2 | 10% |
| **Years of clinical experience** |  |  |
| 1–5 years | 6 | 29% |
| 6–10 years | 4 | 19% |
| 11–20 years | 7 | 33% |
| 21+ years | 4 | 19% |
| **Gender** |  |  |
| Female | 20 | 95% |
| Male | 1 | 5% |
| **Language status** |  |  |
| Monolingual | 10 | 48% |
| Bilingual | 7 | 33% |
| Multilingual (i.e., three or more languages) | 4 | 19% |
| **Language(s) of Service Delivery** |  |  |
| English | 21 | 100% |
| French | 3 | 14% |
| Italian | 1 | 5% |
| Urdu | 1 | 5% |
| Cantonese | 1 | 5% |
| **Race/Ethnicity** |  |  |
| East Asian (e.g. Chinese, Japanese, Korean, Taiwanese descent) | 1 | 5% |
| South Asian (e.g. Indian, Pakistani, Sri Lankan descent) | 4 | 19% |
| White (e.g., European descent) | 15 | 71% |
| Prefer not to answer | 1 | 5% |
| **Practice Setting** |  |  |
| Public Health / Regional Health Authority | 6 | 29% |
| Hospitals (general, paediatric) | 12 | 57% |
| Other (e.g., Community-based services) | 3 | 14% |
| **Community Classification<sup>1</sup>** |  |  |
| Large Urban | 16 | 76% |
| Medium Urban | 2 | 10% |
| Small Urban | 2 | 10% |
| Rural | 1 | 5% |
| **Province** |  |  |
| Ontario | 14 | 67% |
| Alberta | 3 | 14% |
| British Columbia | 1 | 5% |
| Newfoundland & Labrador | 1 | 5% |
| Prince Edward Island | 2 | 10% |

<sup>1</sup>According to Statistics Canada (2022a), areas with a population of at least 1,000 and a density of 400 persons or more per square kilometre are classified as urban population centres, further categorized as small (1,000–29,999), medium (30,000–99,999), or large (100,000+). All other areas are considered rural.

### Domain-Level Summary of Findings

Framework analysis generated 12 themes mapped onto domains of the Theoretical Domains Framework (TDF), illuminating the processes, challenges, and contextual factors influencing clinicians’ collection and use of language history information. Figure 1 presents the relevant TDF domains alongside the emergent themes in relation to the two study research questions. Themes are presented below by theoretical domain, with illustrative quotes attributed using pseudonyms and professional roles.

**Figure 1.**
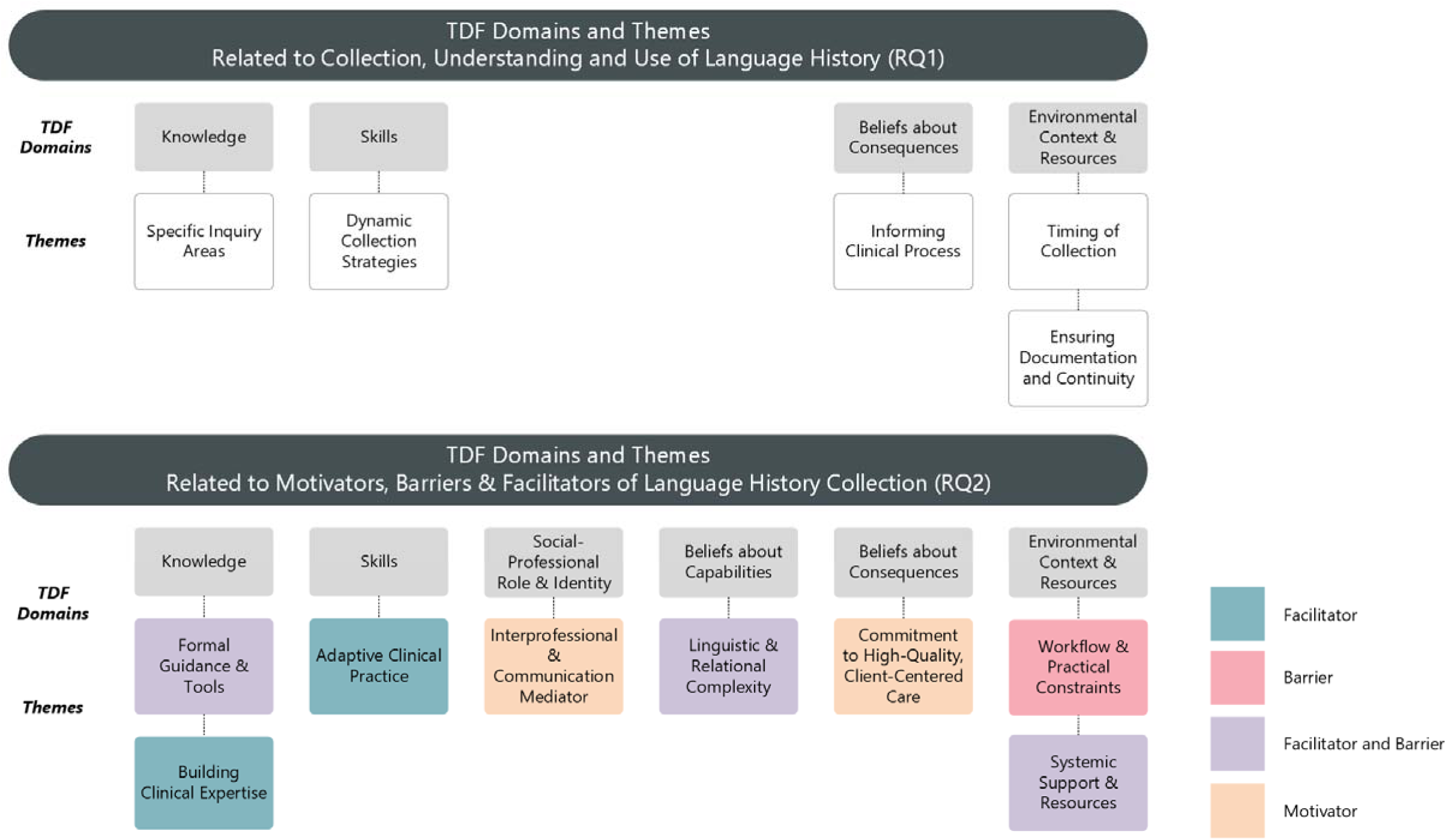
Theoretical Domains Framework (TDF) Domains and corresponding emergent themes related to the collection, understanding, and use of language history (RQ1), and to the motivators, barriers, and facilitators influencing language history collection (RQ2). *Note.* TDF = Theoretical Domains Framework, RQ = Research Question

### Knowledge

#### Specific Inquiry Areas

All clinicians consistently reported collecting a detailed language history, covering a wide range of bilingual factors and related topics, to build a full picture of each child’s language experience. Typically, interviews began by identifying the primary language spoken at home, followed by questions about other languages, who speaks which languages to the child, and in what proportion. Also, where and when languages were used (e.g., home, school, religious or community settings) and age of English exposure was paid attention to especially for newcomers or sequential bilinguals. Assessing language dominance and proficiency was also a priority, including identifying this for different domains. Clinicians also inquired about early milestones, current fluency and literacy and incorporated parents’ perspectives on communication challenges to understand consistency across languages or language-specific difficulties. Language history was often integrated with broader developmental and psychosocial data, including demographics, family structure, medical history, educational placement, community involvement and cultural/immigration background. Clinicians also gathered information about prior services, referral reasons, and practical barriers (e.g., housing, food insecurity), along with preferred language for services, dialects, and interpreter needs to ensure accessible care and contextualization of the child’s needs.

#### Formal Guidance and Tools

A range of formal and self-created tools supported collection during pre-screening and initial assessments. Common resources included “templates,” “screeners,” “case history forms” and “intake interviews” and were viewed generally as facilitators. Nearly all clinicians used setting-specific or custom tools; only one Alberta-based SLP reported frequent use of an externally developed parent interview tool, the Alberta Language and Development Questionnaire (ALDeQ). It was also the only formally developed tool referenced.

A few participants noted limitations in their use of locally developed or clinic-specific forms, including lack of depth. For instance, one participant pointed to a checkbox-based format that restricted elaboration, while others reported that the depth and quality of information varied depending on who completed the referral. It was noted that forms completed by families tended to yield more detailed information, whereas professional referrals often lacked language-related content. Preliminary information could also be gathered by the administrative team and subsequently verified and supplemented by clinicians. No challenges were reported for the ALDeQ.

Knowledge of professional guidelines was mixed. A few participants were unable to name any helpful tools or guidance. A few SLPs referred to specific documents (e.g., Alberta College guidelines, other position statements), while others found existing frameworks too vague to offer actionable structure:

> I guess I tried to piece together from like the position papers on CASLPO [College of Audiologists and Speech-Language Pathologists of Ontario]. I am part of some like SLP forums, and I hear a lot of conversation, and I think more than anything, it’s just made me more sensitized to be really, really careful when assessing a bilingual child. I don’t think that it’s necessarily given me structure on how to do it. (P70, SLP)

#### Building Clinical Expertise

For many, clinical skills evolved with exposure to diverse settings and supervision. Academic training (textbooks, sample templates) provided a starting point, but many felt this was insufficient to feel fully prepared: “You get taught that in school […] but there’s no like scripts that you’re given that you’re expected to follow” (P174, SLP). Peer learning and mentorship were described as key facilitators of skill development, especially for newer clinicians. Informal collaboration was common, and some initiated peer networks when support was lacking. In some settings, mentorship was formally structured through practice leads who observed and provided feedback. Self-initiated learning also played a major role, including engaging with literature, attending seminars, and developing their own tools:

> I’ve probably developed the set of questions that I have in my mind that I’m looking for as a result of reading the literature and getting a sense of what are some of the important factors to consider […] and that’s probably also through attending conferences […] and just kind of picking up that knowledge along the way. (P358, Neuropsychologist)

Access to training varied by setting. One clinician shared that professional development workshops and training modules offered helpful prompts around bilingualism. Others reflected on differences among peers based on experience:

> Some of our older clinicians haven’t maybe kept up with research and some of our newer clinicians haven’t had the same opportunities to learn that I’ve had. […] I don’t know if there is consistency. But in my immediate circles, I feel like we’re on the same page. (P82, SLP)

### Skills

#### Dynamic Collection Strategies

Rather than following rigid intake scripts, many clinicians used a flexible, conversational approach that followed the family’s lead: “it’s more of a conversation than anything […] some of it now is not something I have in writing” (P82, SLP). Some described their process as instinctive:

> I want to say it’s more like an instinct, it just kind of happens as you’re interviewing the family. Like it’s not a protocol or like a list that I’m following that I have in front of me. (P174, SLP)

Clinicians adapted questioning in real time, adjusting the level of detail, reordering topics, “digging” deeper or revisiting earlier points. One clinician described this as “active listening” combined with knowledge. The manner of questioning was also viewed as critical, with clear direct communication prioritized. Inclusive phrasing was used to avoid assumptions: “I don’t want to ask them like, ‘Oh, any languages other than English?’ because I don’t want it to sound like English is […] the be all end all” (P172, Social Worker).

#### Adaptive Clinical Practice

A few clinicians described revising tools and strategies over time to better fit clinical realities by rewording items, updating content and aligning them more closely with population needs: “Our forms are something that we do update fairly often because we want to make them work for us” (P10, SLP). One clinician mentioned revisions to their forms aimed at making language more “equitable or neurodiverse” (P155, SLP).

Many clinicians mentioned that exposure to different cases and support from mentors had helped refine practice. Past cases were described as informing future ones, while remaining open to trial and adjustment. Managing uncertainty was routine; one clinician emphasized developing comfort with ambiguity, asking clarifying questions, and acknowledging the limits of available information:

> Over time, I’ve gotten used to feeling uncomfortable, and knowing that it’s okay to be a bit uncomfortable […] So I think as long as we’re understanding that it’s not perfect, and […] we have to keep working to try to find them. (P61, SLP)

Many clinicians described integrating multiple data sources (interviews, observations, parent forms) and revisiting inconsistencies. Finally, tailoring practices to the specific sociocultural or linguistic background was described as a learned skill: “I try as much as possible to kind of educate myself about how [ESL development] can look. I’ve probably developed the set of questions that I kind of have in my mind as a result of reading the literature” (P358, Neuropsychologist).

### Social-Professional Role & Identity

#### Interprofessional and Communication Mediator

Many SLPs described language history collection as an inherent part of their professional scope. This responsibility was guided by regulatory standards or team protocols. SLPs described an implicit expectation that they would take the lead on language history: “I would say that probably the teams look to the speech pathologists to gather that information” (P61, SLP). This role was also seen as integral to their identity as specialists in language and communication, with SLPs self-describing as often stepping in when other team members lacked professional interest or expertise. SLPs were frequently consulted by colleagues in psychology, occupational therapy, or social work on how to assess or accommodate children’s language needs:

> I’ve been asked, ‘What do I have to think about in terms of language level? […] Can this child express themselves enough in English?’ […] And even the psychologists will sometimes come to us as well looking for guidance, because they don’t love to test with kids who may be new to Canada […] so they will sometimes ask us those questions as well. (P61, SLP)

Non-SLP participants described varied involvement in language history collection: one social worker gathered information to relay to others and sometimes worked interchangeably with other disciplines; a psychologist noted overlapping responsibilities with SLPs.

### Beliefs about Capabilities

#### Linguistic and Relational Complexity

Some clinicians described feeling confident and capable when equipped with relational, communicative skills and extensive relevant experience. For instance, working with younger children whose language histories may follow more predictable patterns was seen as more manageable. Many clinicians linked confidence to the ability to build rapport and read family dynamics. Open-ended questioning was seen as a way to elicit more organic insights. Language skills such as speaking a shared language also acted as a facilitator of rapport and data quality:

> Whenever we have clients come in and they speak Cantonese, for example, they all come to me. […] I find that it’s helpful for families to be able to speak to someone who speaks their language, who understand the child. Because there’s a connection there. If it’s a language that that we understand, I mean, interpreter is good to a point but if you are able to speak the language, I think that would be best. (P69, SLP)

Conversely, linguistic mismatches made the process more taxing.

Other barriers to confidence also emerged. Compared to monolingual patients, bilingualism was seen as more complex, with the added complexity of distinguishing language disorder from language difference: “I find that bilingualism, it’s such a common thing and yet I find it complicated. There’s so much nuance, doing a good assessment and not over-identifying, not under-identifying” (P70, SLP). Confidence was also challenged when information quality was uncertain. Some parents were described as “pretty poor historians” (P166, SLP) and clinicians also expressed scepticism about the objectivity or completeness of parent reports, especially when estimating exposure. Concerns related to stigma and trust also influenced clinicians’ confidence. Some families withheld information about non-English language use, and clinicians worried they might not be hearing the full story but rather what families thought they wanted to hear. Interpreter-mediated communication added another layer of uncertainty. Some noted that interpreters occasionally changed instructions or phrasing, affecting accuracy of interpreted statements. Some families declined interpretation altogether, which raised concerns again about veracity. Others described having no difficulty getting the right information but were less confident in interpreting it:

> I definitely feel fairly confident about collecting information. But where the tricky part comes in is then what to do with it all and what conclusions to make basically. […] In those situations, even though I collect all of this background information, you’re sort of still left with, well, what do I do with the testing that I did? How do I interpret what I did? (P358, Neuropsychologist).

### Beliefs about Consequences

#### Informing the Clinical Process

Many clinicians emphasized language history as core to clinical decision-making, influencing care downstream including assessment, interpretation, planning, and service delivery. One SLP stated, “it’s more like necessity like I don’t think we can do like our due diligence and do our job without knowing that information” (P10, SLP). For children with multilingual backgrounds, even more so: “For me, it’s mandatory for bilingual kids. You can’t have a true picture of a kid if you don’t know what language they’re being exposed to” (P9, SLP).

Language history plays a critical role informs tool selection at the level of assessment:

> It also helps to inform, you know, the approach of assessment and therapy overall because if a child obviously has a stronger language, that’s not English, I’m not going to be using like a standardized test and consulting the norms in English. (P172, SLP)

Many described using language background to contextualize test results, refine diagnostic reasoning, and avoid misdiagnosing language difference as disorder: “Understanding what the child’s exposure to language is like is important because it gives me like a better gauge of is this a true language difference or difficulty versus is this just an exposure concern issue” (P70, SLP). This data also informed service planning and eligibility decisions. As one SLP explained, “The main goal of it is to use the information to proceed with our recommendations, our goals, and any need for further referrals. […] This also helps us determine their eligibility for treatment” (P56, SLP).

#### Commitment to High-Quality, Client-Centered Care

Language history collection was also seen as a means of delivering evidence-based, client-centred care, rooted in professional values and best practice. As one SLP explained,

> If we think of the case history form being kind of like a written extension of our assessment service […] the questions that we’re asking on that form are to gather the information that the research evidence has shown to be necessary to inform practice, like evidence base doesn’t just have to exist for like treatment approaches but there’s evidence base for assessment approaches too. (P10, SLP)

This work was also described as working towards a full picture of the child’s abilities, a best practice in multilingual assessment.

A few emphasized that individualization was key to culturally responsive care: “Just because a family speaks Mandarin doesn’t mean their language practices look like every other Mandarin-speaking family” (P10, SLP). Others linked this understanding directly to goal setting and therapy planning, with no “one size that fits all” (P71, SLP). A few participants linked poor-quality data to poorer outcomes, such as therapy. Many clinicians outside of language-focused roles (e.g., nursing, social work) also emphasized its value for facilitating communication of health information.

### Environmental Context and Resources

#### Timing of Collection

Clinicians described variation in when and how language history was collected, shaped by institutional structures, appointment formats, and case complexity. Collection was often iterative rather than a one-time event, with information gathered pre-visit, during intake, or over follow-up sessions. Institutional structures such as pre-screening teams or intake protocols often influenced how much information was available before the clinician’s direct involvement. However, sometimes this resulted in variability and gaps in information depending on referral pathway. While some teams aimed to gather this information during intake, others approached it as an evolving conversation across sessions. Several clinicians emphasized the importance of verifying and updating information: “Even though the intake form asked what the first language is, I would ask anyway just to double check.” (P56, SLP). These revisits suggest that clinicians often validate and update existing records, rather than treating intake forms as definitive.

Time needed varied widely—from under 5 minutes in straightforward cases to 30–50 minutes in more “complex” ones. For clinicians who integrated language history into a more continuous process, timing was especially difficult to estimate: “It’s probably peppered throughout the whole interview” (P358, Neuropsychologist). Intake forms could sometimes save time as a guide to fill out missing information.

#### Ensuring Documentation and Continuity

Clinicians used a variety of documentation systems to ensure continuity and accessibility of language history across care episodes. These storage options included progress notes (e.g., Subjective, Objective, Assessment, and Plan [SOAP], Data, Action, Response [DAR]), templates, interdisciplinary reports, and in EMRs. Some information, though collected, was not always included in formal documentation unless deemed essential. Clinicians created personal systems and templates for consistency, thoroughness and accuracy:

> I personally use my assessment report template. […] I put all the information that I gathered during the assessment then I go back and refine and make sure that all the information is there and in the right way. (P56, SLP)

Electronic medical records (EMRs) were seen as helpful for documentation of notes and information-sharing. Some platforms even included dedicated language history fields, “There’s also a place on our chart […] that asks what the home language is, what the first language is, and it has a space for comments. […] Everyone can see the information that I’ve gathered” (P82, SLP). However, others noted that it was not often in use or was variable in content.

Team-based care increased the need for clarity and coordination. Clinicians described overlapping efforts across roles: “Tons and tons of my work is with speech paths […] When I read their reports and they read my reports, we’re talking about a lot of the same stuff” (P355, Clinical Psychologist). Others noted differing levels of overlap, again defaulting to more details from those whose scope of practice includes language assessment more explicitly. Accurate documentation also supported long-term care, preparing for reassessments whether it was with the same clinician or another.

#### Workflow & Practical Constraints

Clinicians highlighted time pressure, caseload size, and shared clinical spaces as key constraints. Multidisciplinary assessments with compressed timelines often led to rushed interactions and pressures across roles:

> When the speech language portion takes 40 minutes, I know it puts everybody under pressure. […] what we’ll try to do is while I’m talking to the parent, maybe the OT will come in and get the kid to like walk up the stairs and do some of her motor stuff because it starts to start to feel a little bit of pressure. That’s when things can get missed in terms of case history asking. (P70, SLP)

Dedicated appointments were also constrained by time, as heavy caseloads placed additional pressure on clinicians. These time limitations reduced opportunities to explore alternative tools or approaches:

> We don’t have a whole lot of time to kind of really dibble into […] ‘are there any tools for better understanding how to get a language history?’ because we are so pressed for time, because our caseloads are so heavy […] Our wait lists are so long. (P142, SLP)

Additional practice-related constraints included the need for interpreter services, which were considered essential but introduced added time demands, complexity, and interruptions to the natural flow of conversation. While virtual appointments helped reduce travel time, clinic scheduling pressures, and space limitations, in-person visits remained the preferred approach despite their time-intensive nature. Clinicians also highlighted the considerable administrative burden placed on families, noting, “[It] really is onerous; it’s a lot of asking families to fill in forms, so we actually do our intake appointment in person” (P61, SLP).

#### Systemic Support and Resources

While interpreter services were generally available (e.g., in-person, phone, or audio-visual platforms), they did not always meet the linguistic needs of families, particularly for less common or Indigenous languages. As one nurse described:

> None of the interpretive services have [Low German] because it’s such a specific language. About a year ago now probably, we had problems with the language services line that said it could access Indigenous languages. But we tried to use it for a Cree speaking family. They’re like, ‘What language?’ I say ‘Cree.’ They say, ‘Do you mean Creole?’ I’m like, ‘No, like Cree.’ They’re like, ‘I’ve never heard of this language before. Are you sure this is a language?’ (P391, Nurse)

These gaps, coupled with limited reference materials and insufficient interpreter training for medical contexts, often forced teams to rely on family members or support staff for interpretation, raising concerns about accuracy and appropriateness. This was further exacerbated by staffing shortages and a scarcity of bilingual providers able to support families directly.

While for the most part, workplaces were described as flexible to the clinician practices, one clinician nevertheless expressed frustration with the rigid workflows and system processes that made it difficult to adapt to practice needs:

> We would really like [the hospital] to allow us to send that form through the medical records, so that the parent would be able to fill it out, it would be systematic […] But no, that’s not how we do it yet. (P166, Neuropsychologist)

A few individuals noted that linguistically diverse populations were poorly represented in the research informing clinical tools and system-wide priorities. As one SLP noted, “I think many of us are using a combination of things because nothing ever meets the needs of everybody” (P61, SLP). Another clinician highlighted the need for shared, evidence-based tools accessible across disciplines: “more shared, research-based instruments that can be used across professions […] would strengthen recognition of early language development and support teamwork, because battling silos is a never-ending thing” (P355, Clinical Psychologist).

## Discussion

This study explored how public-sector clinicians collect, interpret, and apply language history information. While recognized as essential to linguistically appropriate and culturally responsive care, language history collection remains underexamined as a discrete clinical task. Our findings reveal it as an intentional and variable practice, shaped by clinician expertise, institutional context and systemic resources. As clinicians’ practices and the factors shaping them were interconnected, the following discussion is broadly organized around our two research questions but incorporates their conceptual overlap.

### Understanding Clinical Practices in Language History Collection

All clinicians in this study universally reported collecting language background information, consistent with high uptake observed among SLPs in diverse cultural and practice settings (Dubasik & Valdivia, 2020; Hallin & Partanen, 2024; Hernandez et al., 2025; Hunt et al., 2025; Selin et al., 2022). Similarly, nearly all clinicians acknowledged the clinical value of language history in guiding assessment and intervention planning, reinforcing its integral role in practice. Clinicians generally collected information about the length and context of first and second language exposure (e.g., home versus school) and estimated language proficiency, alongside developmental, psychosocial, and practical considerations – embedding language history within a holistic, family-centred assessment.

The linguistic variables (e.g., age and mode of exposure) that clinicians focused on have been shown to determine the language abilities of bilingual children (Kohnert, 2010; McLeod et al., 2017; Paradis et al., 2010). In this regard, clinicians tended to collect language background information consistent with the research evidence, which should be helpful in determining a linguistic profile. However, there was still considerable variability in the types and depth of information collected, how and when the information is collected and how it is used to inform clinical decisions. Language history collection occurred across different points in the care pathway, across intake, assessment and at follow up. This iterative practice allowed for clarification and responsiveness to family circumstances but also risked incomplete records, and potentially misguided assessment and intervention planning. Documentation methods also varied, with information being recorded in electronic medical record fields, reports and notes. These differences significantly shaped what information was gathered, how it was recorded, and how easily it could be shared across clinical teams.

Most clinicians relied on locally developed forms, templates, and interviews reflecting an adoption of dynamic, adaptable, context-specific approaches. Many preferred conversational interviews that allowed them to follow the family’s lead and tailor questions in real time. This method was often perceived as more effective, especially when their other options did not reflect the family’s language practices or were overly restrictive (e.g., design elements such as checkboxes). However, clinicians also expressed uncertainty about whether they were collecting enough or the right type of information and also doubted the veracity due to the information source (e.g., family members, other clinicians, non-clinical staff). While adaptability is a strength in complex care environments, it may also contribute to inconsistencies in the quality, completeness, and application of language history data, especially in the absence of formal guidance or structure. These varied approaches of more structured versus flexible methods present with potential benefits and drawbacks, and yet no research has directly compared their clinical impact.

SLPs were typically regarded as the professionals leading the collection of language history, reflecting their disciplinary expertise in language and communication. This mirrored previous descriptions of SLPs as language specialists in multidisciplinary contexts (Pagan et al., 2015; Sander et al., 2009), with other clinicians also having intersecting roles and participating in language history collection as well. These findings reinforce the need for shared frameworks and interprofessional coordination, a priority echoed by participants who supported the development of research-based tools to reduce duplication and promote consistency across roles.

### Barriers, Supports, and Decision-Making in Practice

Clinicians’ motivations for collecting language history were clear: language history was seen as essential to accurate assessment, equitable service planning, and culturally responsive care. However, numerous systemic, professional, and contextual factors influenced its implementation.

One prominent influence on clinician practice was the lack of formal training and consistent guidance. While some clinicians referenced provincial guidelines or professional position statements, many were unaware of formal resources or found them too vague to offer actionable support. In the absence of structured support, clinicians relied on peer exchange, clinical judgment, and informal mentorship and supplemental learning opportunities. This variability raised concerns about inconsistencies in practice, especially across disciplines or among clinicians with differing levels of experience.

At the same time, this variability appeared to foster the development of adaptive expertise, the capacity to respond flexibly to clinical uncertainty and contextual variability (Mylopoulos & Woods, 2017). Clinicians described adjusting their questioning strategies in response to family narratives, clinical concerns and evolving circumstances. Some revised their intake tools over time to reflect population needs; others constructed mental checklists based on literature or case experience. In many cases, this meant constructing their own frameworks in the absence of formal tools or relying on conversational interviews when time or interpreter constraints precluded more structured methods. Rather than indicating a lack of knowledge, these practices often reflected skilled improvisation grounded in professional experience.

Still, adaptive expertise does not eliminate the risks associated with unstructured practice. Some clinicians expressed doubt about the accuracy or sufficiency of the information gathered, for instance when language interpretation was unreliable. Others described feeling confident in data collection, but less so in data interpretation, especially when deciding how language background might affect testing or diagnostic reasoning. These challenges suggest that while adaptive expertise is a strength, it needs to be supported by structured resources and professional development but not used as a substitute for them.

Time pressures, high caseloads, limited appointment durations and competing demands were frequently cited as major constraints. Clinicians often had to prioritize immediate concerns, limiting the depth of language inquiry. These constraints often led clinicians to prioritize immediate concerns over in-depth exploration of linguistic background. Documentation systems further restricted practice—some EMRs lacked structured fields for language history, making it difficult to ensure visibility across teams. Interpreter access also shaped clinicians’ ability to collect meaningful language histories.

At the organizational level, institutional policies influence flexibility in workflow. Some settings permitted clinicians to adapt tools and forms to match the needs of multilingual families; others required rigid adherence to established processes. This variation significantly impacted the feasibility of implementing more nuanced or responsive approaches to lessen the potential burden that language history could place on the family and clinician. Clinicians also noted that linguistically diverse populations were often underrepresented in research and tool development, limiting the applicability of research-based instruments.

Although several structured and evidence-informed tools exist to support language history collection—like the Alberta Language and Development Questionnaire (ALDeQ; Paradis et al., 2010), Quantifying Bilingual Exposure questionnaire (Q-BEx; De Cat et al., 2023), Bilingual Input-Output Survey (BIOS; Peña et al., 2018), or the LEAP-Q (Marian et al., 2007) – few participants were familiar with or regularly used them. Some clinicians expressed concerns that they lacked training or time to better understand these options. Others found them too rigid or not adaptable enough to the family’s sociolinguistic context. Yet interestingly, several clinicians described tailoring their questions based on emerging concerns, an approach that mirrors the modular, flexible design of tools like Q-BEx. This suggests that the problem may not be the tools themselves, but their fit with clinical workflows, lack of training, or insufficient awareness among practitioners. Effective use of these tools may require theoretical knowledge or training not commonly available, in time-pressed or resource-limited clinical environments (Cunningham et al., 2017; Thome et al., 2020). Reviews such as Leung and Molnar’s (2025) confirm this implementation gap: of seven identified child language history tools, only two demonstrated both developmental rigor and pragmatic utility. Although a few clinical adaptations have been validated, uptake remains low in practice. These findings suggest that tool design alone is insufficient. High-quality instruments must also be based on stakeholder consultation, integrated into workflow, supported by training, and made accessible through documentation systems and team processes.

### Bridging the Gap: Practical Paths Forward

The tension between flexibility and structure emerged as a central theme across both research questions. Clinicians valued the ability to adapt their questioning to the family’s context. These practices demonstrated clinical insight, cultural humility, and responsive care. Some also recognized the limitations of relying solely on these more flexible, unstructured methods. This trade-off was particularly evident when clinicians felt unsure about the completeness or reliability of the information they had gathered, especially in the absence of formal tools or trained interpreters.

A clear opportunity exists for hybrid approaches that preserve the strengths of adaptive expertise while providing structured support. There is a growing need for hybrid approaches that balance evidence-based structure with clinician flexibility. Tools should be pragmatic, brief, flexible, and interprofessional, while also being embedded in workflows and supported through training (Glasgow & Riley, 2013), with the Q-BEx and ALDeQ as examples. Beyond tools, shared protocols and interprofessional coordination are essential to ensure consistency and reduce duplication across roles.

Participants emphasized the need for system-wide supports: documentation protocols and tools, robust interpreter services, culturally responsive intake processes, and institutional flexibility. Language history collection is not simply a means to collect background information, it is a foundational component of equitable, evidence-based clinical care. Ensuring its quality and consistency requires not only clinician initiative, but leadership at the institutional, policy, and professional levels.

### Limitations

As our recruitment materials highlighted multilingualism, self-selection bias for clinicians with greater interest or confidence in this area may have been introduced. Perspectives from non-clinical roles were not included, which may limit the generalizability of findings across all relevant roles. However, the study deliberately prioritized professionals most closely involved in the language history collection process.

While the TDF provided a structured lens to examine clinicians’ beliefs and behaviours, its focus on individual-level factors limited exploration of broader systemic and relational influences, such as institutional policies or interprofessional dynamics. To address this, we used a combined inductive and deductive approach, allowing themes to emerge both from the data and in alignment with the TDF. As with all qualitative research, findings reflect participants situated perspectives and the interpretive lens of the research team.

## Conclusion

Language history collection in public-sector healthcare is a complex, multidimensional practice shaped by clinician initiative, institutional structures, and systemic resources. Far from being a routine intake step, it is central to equitable and accurate assessment, requiring both specialized expertise and supportive systems. Strengthening tools, training, and interprofessional collaboration will be key to ensuring that this essential component of culturally responsive care is delivered consistently and effectively across settings. Ongoing investment in hybrid tools, integrated training, and systems-level coordination will be critical to closing the gap between policy and practice in multilingual care delivery.

## Acknowledgments

The authors extend their sincere gratitude to all participants who generously shared their time and experiences, making this study possible. We also gratefully acknowledge the support of research assistants Dr. Glynnis Dubois and Victoria Gotcheva for their invaluable assistance with transcription and data verification.

## Data Availability Statement

Due to the qualitative nature of this research and to protect participant confidentiality, the interview transcripts and related data are not publicly available. Supplementary materials related to the study are available with this article.

## Ethics Approval Statement

This study was reviewed and approved by the University of Toronto Research Ethics Board (Protocol #46019). All participants provided informed consent prior to participation.

## References

American Psychological Association. (2020). APA guidelines for psychological assessment and evaluation. In American Psychological Association: Washington, DC, USA.

American Speech-Language-Hearing Association. (n.d.). Multilingual service delivery in audiology and speech-language pathology [Practice Portal]. Retrieved October 7, 2024, from https://www.asha.org/Practice-Portal/Professional-Issues/Bilingual-Service-Delivery/

American Speech-Language-Hearing Association. (2023). Ad Hoc Committee on Bilingual Service Delivery: Competencies, expectations, and recommendations for multilingual service delivery. https://www.asha.org/siteassets/reports/ahc-bilingual-service-delivery-final-report.pdf

American Speech-Language-Hearing Association. (2025). 2024 Member and Affiliate Profile. https://www.asha.org/siteassets/surveys/2024-member-affiliate-profile.pdf

Canadian Association of Social Workers. (2024). Social work and primary care: A vision for the path forward. Canadian Association of Social Workers. https://www.casw-acts.ca/files/attachements/4023_CASW_SocialWorkAndPrimaryCare_Report_EN-Final.pdf

Cane, J., O’Connor, D., & Michie, S. (2012). Validation of the theoretical domains framework for use in behaviour change and implementation research. Implementation Science, 7(1), 37. 10.1186/1748-5908-7-37

Cao, X., & Keller-Bell, Y. (2025). Assessing Speech-Language Pathology Graduate Student Clinicians’ Services With Culturally and Linguistically Diverse Clients. Perspectives of the ASHA Special Interest Groups, 10(3), 1042–1053. 10.1044/2025_PERSP-24-00242

Cunningham, B. J., Hidecker, M. J. C., Thomas-Stonell, N., & Rosenbaum, P. (2017). Moving research tools into practice: The successes and challenges in promoting uptake of classification tools. Disability and Rehabilitation, 40(9), 1099–1107. 10.1080/09638288.2017.1280544

De Cat, C., Kašćelan, D., Prévost, P., Serratrice, L., Tuller, L., & Unsworth, S. (2023). How to quantify bilingual experience? Findings from a Delphi consensus survey. Bilingualism, 26(1). 10.1017/S1366728922000359

Douglas, M. K., Rosenkoetter, M., Pacquiao, D. F., Callister, L. C., Hattar-Pollara, M., Lauderdale, J., Milstead, J., Nardi, D., & Purnell, L. (2014). Guidelines for Implementing Culturally Competent Nursing Care. Journal of Transcultural Nursing, 25(2), 109–121. 10.1177/1043659614520998

Dröge, K. (2024). noScribe. AI-powered Audio Transcription (Version 0.5) [Computer software]. https://github.com/kaixxx/noScribe

Dubasik, V. L., & Valdivia, D. S. (2020). School-Based Speech-Language Pathologists’ Adherence to Practice Guidelines for Assessment of English Learners. Language, Speech, and Hearing Services in Schools, 52(2), 485–496. 10.1044/2020_LSHSS-20-00037

Fujii, D. E. M. (2018). Developing a cultural context for conducting a neuropsychological evaluation with a culturally diverse client: The ECLECTIC framework*. Clinical Neuropsychologist, 32(8). 10.1080/13854046.2018.1435826

Fulcher-Rood, K., & Castilla-Earls, A. (2023). Differences in Child Language Assessment Practices Between School-Based and Non–School-Based Speech-Language Pathologists: Results From a Nationwide Survey. Language, Speech, and Hearing Services in Schools, 54(4), 1117–1135. 10.1044/2023_LSHSS-22-00185

Fumero, K., Torres-Chavarro, M., & Wood, C. (2021). Challenges in Service Provision to Children and Families from Linguistically Diverse Backgrounds. Seminars in Speech and Language, 42, 395–418. 10.1055/s-0041-1736498

Gale, N. K., Heath, G., Cameron, E., Rashid, S., & Redwood, S. (2013). Using the framework method for the analysis of qualitative data in multi-disciplinary health research. BMC Medical Research Methodology, 13(1), 117. 10.1186/1471-2288-13-117

Glasgow, R. E., & Riley, W. T. (2013). Pragmatic Measures: What They Are and Why We Need Them. American Journal of Preventive Medicine, 45(2), 237–243. 10.1016/J.AMEPRE.2013.03.010

Guiberson, M., & Atkins, J. (2012). Speech-Language Pathologists’ Preparation, Practices, and Perspectives on Serving Culturally and Linguistically Diverse Children. Communication Disorders Quarterly, 33(3), 169–180. 10.1177/1525740110384132

Hallin, A. E., & Partanen, P. (2024). Factors affecting speech-language pathologists’ language assessment procedures and tools – challenges and future directions in Sweden. Logopedics Phoniatrics Vocology, 49(3), 104–113. 10.1080/14015439.2022.2158218

Harris, P. A., Taylor, R., Minor, B. L., Elliott, V., Fernandez, M., O’Neal, L., McLeod, L., Delacqua, G., Delacqua, F., Kirby, J., & Duda, S. N. (2019). The REDCap consortium: Building an international community of software platform partners. Journal of Biomedical Informatics, 95, 103208. 10.1016/j.jbi.2019.103208

Harris, P. A., Taylor, R., Thielke, R., Payne, J., Gonzalez, N., & Conde, J. G. (2009). Research electronic data capture (REDCap)—A metadata-driven methodology and workflow process for providing translational research informatics support. Journal of Biomedical Informatics, 42(2), 377–381. 10.1016/j.jbi.2008.08.010

Hernandez, M., Fulcher-Rood, K., & Castilla-Earls, A. (2025). Speech-Language Pathologists’ Perspectives on Language Assessment in Bilingual Children. Language, Speech, and Hearing Services in Schools, 1–20. 10.1044/2025_LSHSS-24-00145

Hunt, E. F., Nang, C., Meldrum, J. S., & Armstrong, E. (2025). Assessment of multilingual children by Western Australian speech-language pathologists: A survey of practices, barriers and facilitators. International Journal of Speech-Language Pathology, 27(3), 370–384. 10.1080/17549507.2025.2499516

International Expert Panel on Multilingual Children’s Speech. (2012). Multilingual children with speech sound disorders: Position paper. Research Institute for Professional Practice, Learning & Education (RIPPLE), Charles Sturt University. http://www.csu.edu.au/research/multilingual-speech/position-paper

Kašćelan, D., Prévost, P., Serratrice, L., Tuller, L., Unsworth, S., & De Cat, C. (2022). A review of questionnaires quantifying bilingual experience in children: Do they document the same constructs? Bilingualism, 25(1). 10.1017/S1366728921000390

Kohnert, K. (2010). Bilingual children with primary language impairment: Issues, evidence and implications for clinical actions. Journal of Communication Disorders, 43(6), 456–473. 10.1016/J.JCOMDIS.2010.02.002

Kritikos, E. P. (2003). Speech-Language Pathologists’ Beliefs About Language Assessment of Bilingual/Bicultural Individuals. American Journal of Speech-Language Pathology, 12(1), 73–91. 10.1044/1058-0360(2003/054)

Lal, P. B., Wishart, L. R., Ward, E. C., Schwarz, M., Seabrook, M., & Coccetti, A. (2023). Understanding barriers and facilitators to speech-language pathology service delivery in the emergency department. International Journal of Speech-Language Pathology, 25(4), 509–522. 10.1080/17549507.2022.2071465

Leung, K. I., & Molnar, M. (2025). Bilingualism measures in children: A critical review of content overlap, development, and pragmatic quality. International Journal of Speech-Language Pathology. 10.1080/17549507.2025.2532787

Lincoln, Y. S., & Guba, E. G. (1986). But is it rigorous? Trustworthiness and authenticity in naturalistic evaluation. New Directions for Program Evaluation, 1986(30), 73–84.

Lumivero. (2023). *NVivo* (Version 14) [Computer software].

McKenna, M., Soto-Boykin, X., Larson, A., & Julbe-Delgado, D. (2025). Speech-Language Therapists’ Training, Confidence, and Barriers When Serving Bilingual Children: Development and Application of a National Survey. American Journal of Speech-Language Pathology, 1–17. 10.1044/2025_ajslp-24-00498

McLeod, S., Verdon, S., & The International Expert Panel on Multilingual Children’s Speech. (2017). Tutorial: Speech assessment for multilingual children who do not speak the same language(s) as the speech-language pathologist. American Journal of Speech-Language Pathology, 26(3). 10.1044/2017_AJSLP-15-0161

Melo, A. T. S. de, Barbosa, G. D., Jesus, E. M. S. de, Matos, A. L. dos S., Santos, E. M. de S., Barreto, Í. D. C., Alves, M. V. M., & Medeiros, A. M. C. (2022). Clinical history speech-language pathology protocols: Integrative review. Audiology - Communication Research, 27, e2673. 10.1590/2317-6431-2022-2673en

Morgan, D. L. (2014). Pragmatism as a Paradigm for Social Research. Qualitative Inquiry, 20(8), 1045–1053. 10.1177/1077800413513733

Multilingual-Multicultural Affairs Committee, International Association of Communication Sciences and Disorders. (2011). Recommendations for Working with Bilingual Children.

Multilingual-Multicultural Affairs Committee, International Association of Communication Sciences and Disorders. (2020). Common Questions by Speech Language Therapists / Pathologists about Bilingual / Multilingual Children and Informed, Evidence-based Answers. https://ialpglobal.org/wp-content/uploads/2025/02/FAQs-for-Speech-and-Language-Therapists-Pathologists_ENGLISH.pdf

Mylopoulos, M., & Woods, N. N. (2017). When I say… Adaptive expertise. Medical Education, 51(7). 10.1111/medu.13247

NoorAli, S., De Anda, S., Cycyk, L. M., & Starlin, S. (2025). Barriers and Facilitators to Assessment Practices in Linguistically Diverse Children: A Preliminary Application of Theoretical Domains Framework. American Journal of Speech-Language Pathology, 1–22. 10.1044/2024_AJSLP-24-00256

Onwuegbuzie, A. J., & Leech, N. L. (2005). On Becoming a Pragmatic Researcher: The Importance of Combining Quantitative and Qualitative Research Methodologies. International Journal of Social Research Methodology, 8(5), 375–387. 10.1080/13645570500402447

Pagan, E., Ownsworth, T., McDonald, S., Fleming, J., Honan, C., & Togher, L. (2015). A survey of multidisciplinary clinicians working in rehabilitation for people with traumatic brain injury. Brain Impairment, 16(3), 173–195.

Paradis, J. (2011). Individual differences in child English second language acquisition: Comparing child-internal and child-external factors. Linguistic Approaches to Bilingualism, 1(3), 213–237. 10.1075/LAB.1.3.01PAR

Paradis, J., Emmerzael, K., & Duncan, T. S. (2010). Assessment of English language learners: Using parent report on first language development. Journal of Communication Disorders, 43(6), 474–497. 10.1016/J.JCOMDIS.2010.01.002

Parveen, S., & Santhanam, S. P. (2021). Speech-Language Pathologists’ Perceived Competence in Working With Culturally and Linguistically Diverse Clients in the United States. Communication Disorders Quarterly, 42(3), 166–176. 10.1177/1525740120915205

Peña, E. D., Gutiérrez-Clellen, V. F., Iglesias, A., Goldstein, B. A., & Bedore, L. M. (2018). Bilingual English Spanish Assessment (BESA). Brookes.

Ritchie, J., Spencer, L., Bryman, A., & Burgess, R. G. (1994). Analyzing qualitative data.

Royal College of Speech and Language Therapists. (2006). Communicating quality 3: RCSLT’s guidance on best practice in service organisation and provision. Royal College of Speech and Language Therapists.

Sander, A. M., Raymer, A., Wertheimer, J., & Paul, D. (2009). Perceived Roles and Collaboration Between Neuropsychologists and Speech-Language Pathologists in Rehabilitation. The Clinical Neuropsychologist, 23(7), 1196–1212. 10.1080/13854040902845706

Santhanam, S. P., Gilbert, C. L., & Parveen, S. (2019). Speech-Language Pathologists’ Use of Language Interpreters With Linguistically Diverse Clients: A Nationwide Survey Study. Communication Disorders Quarterly, 40(3), 131–141. 10.1177/1525740118779975

Selin, C. M., Rice, M. L., Girolamo, T. M., & Wang, C. J. (2022). Work Setting Effects on Speech-Language Pathology Practice: Implications for Identification of Children With Specific Language Impairment. American Journal of Speech-Language Pathology, 31(2), 854–880. 10.1044/2021_AJSLP-21-00024

Shipley, K. G., & McAfee, J. G. (2023). Assessment in Speech-Language Pathology: A Resource Manual, Seventh Edition. Plural Publishing.

Speech Pathology Australia. (2023). Working in a Culturally and Linguistically Diverse Society—Clinical Guideline. https://www.speechpathologyaustralia.org.au/resource?resource=119

Speech-Language and Audiology Canada. (2018). SAC Salary & Benefits Survey 2018: Compensation Analysis of Speech-Language Pathologists in Canada (pp. 1–46). https://www.sac-oac.ca/practice-resources/resource-library/salary-benefits-survey-reports/

Speech-Language and Audiology Canada. (2024). SAC Position Paper on School-Based Speech-Language Pathology Services in the Context of Multilingualism.

Statistics Canada. (2022a). Dictionary, Census of Population, 2021: Population centre (POPCTR). https://www12.statcan.gc.ca/census-recensement/2021/ref/dict/az/Definition-eng.cfm?ID=geo049a

Statistics Canada. (2022b). While English and French are still the main languages spoken in Canada, the country’s linguistic diversity continues to grow (The Daily). https://www150.statcan.gc.ca/n1/daily-quotidien/220817/dq220817a-eng.htm

Suswaram, S., Brady, N. C., & Gillispie, M. (2023). The role of service providers’ linguistic backgrounds on assessment of multilingual children. Journal of Communication Disorders, 102, 106302. 10.1016/j.jcomdis.2023.106302

Teoh, W. Q., Brebner, C., & McAllister, S. (2018). Bilingual assessment practices: Challenges faced by speech-language pathologists working with a predominantly bilingual population. *Speech*, Language and Hearing, 21(1), 10–21. 10.1080/2050571X.2017.1309788

Thome, E. K., Loveall, S. J., & Henderson, D. E. (2020). A Survey of Speech-Language Pathologists’ Understanding and Reported Use of Evidence-Based Practice. Perspectives of the ASHA Special Interest Groups, 5(4), 984–999. 10.1044/2020_PERSP-20-00008

Tong, A., Sainsbury, P., & Craig, J. (2007). Consolidated criteria for reporting qualitative research (COREQ): A 32-item checklist for interviews and focus groups. International Journal for Quality in Health Care, 19(6), 349–357. 10.1093/intqhc/mzm042

Verdon, S., McLeod, S., & Wong, S. (2015). Supporting culturally and linguistically diverse children with speech, language and communication needs: Overarching principles, individual approaches. Journal of Communication Disorders, 58, 74–90. 10.1016/j.jcomdis.2015.10.002

Williams, C. J., & McLeod, S. (2012). Speech-language pathologists’ assessment and intervention practices with multilingual children. International Journal of Speech-Language Pathology, 14(3), 292–305. 10.3109/17549507.2011.636071

